# Palliative Care in Geriatric Trauma: Quantitative Insight From 64 Trauma Surgeon Survey Respondent on Utilizing Specialty Palliative Care in Geriatric Trauma

**DOI:** 10.1101/2024.12.11.24318848

**Authors:** Morgan J. Hopp, Cameron E. Comrie, Paul T Kang, Jacob J. Strand, Wil L. Santivasi, Alexzandra K. Hollingworth, Gaby Iskander, Jordan Weinberg, Kelly L. Wu

## Abstract

**Background:** Early specialty palliative care (SPC) involvement in geriatric trauma care improves outcomes, quality of life and healthcare utilization. However, SPC usage is inconsistent and imprecise. A knowledge gap persists in understanding surgeons’ perspectives towards SPC and barriers in geriatric trauma.

**Methods:** The 38-question survey was distributed through a prestigious surgical society’s membership. Subsequently, comparative analysis of responses was completed based on demographic features.

**Results:** 64 surgeons responded (2.8%). 87.5% of respondents identified a potentially life-limiting diagnosis/prognosis and 76.6% conflicting goals of care as consult triggers. 59.4% reported comfort in addressing the palliative needs without SPC consult. The perception of limited SPC availability (54.7%) was a common barrier. 28.1% felt that patient/family resistance was the most common reason not to consult SPC.

**Conclusions:** Surgeons reported comfort with goals of care discussions, perceived limited SPC availability, and the perception of patient/family resistance as limitation to consultation. These data provide previously unexplored insight from trauma surgeons.

## Background

Since 2010, there has been a 34.2% increase in the number of Americans over 65 years old ^1^. The increase in the national geriatric population is multi-faceted, with one of the most significant factors being the effect of geriatric-focused medicine on improving life expectancy^2^. Compared to younger adults with similar injuries, elderly trauma patients have increased risks of morbidity, mortality, and functional decline ^3–5^. Geriatric patients’ lower cardiac output, increase in pulmonary complications, and delirium place them at increased risk for surgical complications ^6–9^. Adequate care for the geriatric population after trauma requires specialized services, including but not limited to pharmacists with expertise in geriatric polypharmacy management, geriatricians, case managers, expedited care in the emergency department, and expedited time to the operating room ^4,10^.

In 2016, patients 55 years and older accounted for over 40% of all traumas and over 55% of all trauma-related deaths ^11^. Geriatric trauma patients incur a disproportionate amount of the total US trauma expenses due to the complexities of geriatric care^12^. Many efforts have been made to identify best practices for geriatric surgical patients. The American College of Surgeons established the Geriatric Verification Program to optimize the surgical care of aging adults ^10,13^. In recent years, a growing body of research by trauma surgeons and trauma nurses has looked at the benefits of Specialty Palliative Care (SPC) in the outcomes of geriatric trauma patients ^3–5,12,14–16^. SPC is “is specialized medical care for people living with a serious illness, focused on providing patients with relief from the symptoms and stress of the illness” ^17^ also described as “multidisciplinary specialty that aims to relieve suffering and support quality of life for seriously ill patients and their families” ^18^. It can be summarized into three focuses: (1) symptom management, (2) psychosocial support and (3) aiding with decision-making. In geriatric trauma patients, early use of SPC is associated with improved symptom management, reduced length of stay, increased discharge to hospice, and reduced healthcare utilization at the end-of-life ^19,20^. In seriously ill older adults hospitalized for heart failure and malignancy inpatient, SPC consultation is associated with improvements in pain, depression, functional decline, and caregiver burden ^12,14,16,21–23^. However, current literature suggests that over half of older trauma patients have unmet SPC needs at the time of discharge ^24,25^, and that SPC is underutilized in older adults admitted for trauma ^24,26^. Across many fields of medicine, studies have shown that SPC is delivered late in the patient’s course or upon uncontrolled symptoms, which diminishes the potential effectiveness of the consult on quality of life ^21,27–30^. When used promptly, SPC consultation improves the quality of care and results in a decrease in goal-discordant care^31,32^. Based on the results of studies in various fields of medicine ^12,14,16,27–32^, the early and effective involvement of SPC in the treatment of geriatric trauma patients has the potential to directly improve patient outcomes, quality of life and patient-centered approaches to care.

To the best of our knowledge, no qualitative or quantitative study of any size, has investigated the barriers and decision-making by which trauma surgeons consult SPC for the management of geriatric trauma patients. This single time point study aimed to initiated insight into the current utilization of SPC consult services in geriatric trauma patients and the pertinent barriers from the perspective of trauma surgeons survey respondents.

## Methods

### Survey Respondents

A national cross-sectional survey among trauma providers was conducted. The survey was sponsored by the American Association for the Surgery of Trauma (AAST) and was approved by Dignity Health St. Joseph’s Medical Center’s Institutional Review Board (IRB# PHXNR-23-500-253-71-47). It was developed and disseminated using Redcap (Nashville, Tennessee, USA), an online secure data collection tool and distribution software.

The AAST email list was used as a convenient sample. Participants were invited to complete the survey using an anonymous, electronic link sent via the AAST listserv from June through August 2023. Survey recruitments were sent as separate emails every 2 weeks during the three months (a total of 6 emails) and embedded in the weekly/monthly newsletter during this time. The survey link was also available to be accessed via the AAST webpage. Participation in the survey was voluntary and respondents consented to participate by completing and submitting the survey. Incomplete responses were excluded from the analysis of each individual question. The survey was estimated to take 5–10□min to complete.

### Survey Instrument

Two trauma physicians (JW, AH) a Palliative Care physician (KW), and one medical student (MH) initially developed the survey items based on guideline recommendations and gaps. A trauma physician (HS-L) and a medical librarian (KK) who were not study investigators completed pilot testing to review content, determine ease of use, and identify errors. Additionally, the survey underwent review by the AAST Surveys Subcommittee composed of four trauma physicians. Feedback from reviewers was then incorporated into the final version and the changes were approved by study investigators.

The survey was a 38-item questionnaire. It was separated into: Demographics, Decision-Making Insight, and Palliative Care Utilization. All sections consisted of multiple-choice questions with single or multiple responses as appropriate. The likert scale was only utilized in the demographic question regarding frequency of SPC consultation in geriatric trauma patients. Display logic was used for 17 questions based on previous responses. For example, if respondents answered that the trauma service at the hospital where they primarily work acts as a consulting service rather than the admitting service, the next question asked which specialty acts as the admitting service. All questions with “other” option had a subsequent optional free text box allowing for explanation (Supplemental Table 1).

### Statistical Analysis

All variables for this analysis were categorical or ordinal variables, thus descriptive statistics were reported as frequencies and percentages for all categories. Two-sample comparisons of responses based on demographic factors (trauma center type, trauma level designation, board certification, years in practice, practice setting, percentage of geriatric patients in the practice setting, and amount of palliative medicine consults) were done using the Wilcoxon Rank Sum Test for ordinal variables and Chi-squared or Fisher’s Exact Test for categorical variables. Comparisons between three or more groups were conducted using the Kruskal-Wallis Test for ordinal variables. All p-values were two-sided and p<0.05 was considered statistically significant. All data analyses were conducted using STATA version 18 (StataCorp; College Station, TX).

## Results

### Demographics

Sixty-four out of 2255 AAST members responded and were included in the analysis, a response rate of 2.8%. Respondent demographics are reported in Table 1. Respondents were 40.6% female and 57.8% male. AAST members at all career stages participated from those currently in residency or fellowship (7/64;10.9%) to trauma physicians practicing over 20 years (18/64;28.1%). Most respondents were at level 1 trauma centers (50/65; 78.1%) with 61 respondents who identified the admitting services at their institution, all identified trauma as the admitting service (3 missing data). 52 worked at trauma centers with a subspecialty SPC consult team (52/64; 81.3%). Additionally, a majority of respondents estimated that 25-50% of their patients were geriatric (31/64; 48.4%), and 25 estimated that over 50% of their patient population were geriatric (25/62; 40.3%).

**Table 1:** Demographics of Respondents.

| Survey Item | N (%) |
| --- | --- |
| Gender |  |
| Female | 26 (40.6%) |
| Male | 37 (57.8%) |
| Board Certifications – Select all that Apply |  |
| General surgery | 54 (84.4%) |
| Critical care | 48 (75.0%) |
| Trauma and acute care surgery | 22 (34.4%) |
| Neuro critical care | 5 (7.8%) |
| Hospice and palliative medicine | 1 (1.6%) |
| Other | 2 (3.1%) |
| How many years have you been a practicing attending physician? |  |
| Currently in residency/fellowship | 7 (10.9%) |
| Under 5 | 14 (21.9%) |
| 6-10 | 11 (17.2%) |
| 11-15 | 5 (7.81%) |
| 16-20 | 6 (9.38%) |
| 21+ | 18 (28.1%) |
| How would you describe your primary practice setting? |  |
| Academic | 44 (68.8%) |
| Urban | 36 (56.3%) |
| Community | 13 (13.3%) |
| Rural | 2 (3.1%) |
| Estimate the percentage of geriatric (60+ years old) trauma patients in your current practice. |  |
| Less than 25% | 6 (9.4%) |
| 25-50% | 31 (48.4%) |
| 51-75% | 19 (29.7%) |
| Over 75% | 6 (9.4%) |
| How often do you consult Palliative Care in geriatric (60+ years old) trauma patients? |  |
| Rarely | 6 (9.4%) |
| Sometimes | 25 (39.1%) |
| Often | 24 (37.5%) |
| Always | 1 (1.6%) |
| What level trauma center do you primarily work at? |  |
| Level 1 | 50 (78.1%) |
| Level 2 | 9 (14.1%) |
| Level 3 | 1 (1.6%) |
| Other | 4 (6.3%) |
| The trauma service at the hospital that I primarily work is a/an: |  |
| Admitting service | 61 (95.3%) |
| Which specialty acts as the admitting service? |  |
| IM/Hospitalist | 0 (0.0%) |
| General Surgery | 0 (0.0%) |
| Critical Care (IM) | 0 (0.0%) |
| Critical Care (Surgery) | 0 (0.0%) |
| Other | 0 (0.0%) |
| Does your trauma center have a physician run subspecialty Palliative Care consult team? |  |
| Yes | 52 (81.3) |
| No | 12 (19.7) |
| How often do you consult Palliative Care in geriatric (60+ years old) trauma patients? |  |
| Rarely | 6 (9.38) |
| Sometimes | 25 (39.1) |
| Often | 24 (37.5) |
| Always | 1 (1.56) |
| Missing | 8 (12.5) |

### Decision-Making Insights

Examining decision-making insights (Table 2), 23/64 (35.9%) responded that their trauma center had a specialized geriatric service. Potentially life-limiting diagnosis/poor prognosis was the most identified factor for prompting an SPC consult (56/64; 87.5%), followed by conflicting goals of care (49/64; 76.6%) and when different members of the team (staff/patients/family), do not agree on the plan of care (31/64; 48.4%). When considering age as a factor for consulting SPC, the majority identified 70 years and older as the age group of most concern (Table 2). 89.1% (57/64) identified the trauma surgeon as the team member who initiates the need for SPC consultation, followed by the resident and advanced practitioner. Respondents reported that the top three benefits of consulting SPC were support with goals of care or shared medical decision-making (81.3%), added support for patients and families (79.7%), and improved communication between staff and family (67.2%) (Table 2). The most common barrier to consulting SPC was the perception of limited staffing/availability of the SPC team (54.7%) (Table 2). 59.4% reported that the most common reason not to consult SPC is that they, the responding trauma surgeon, feel comfortable addressing the palliative needs of the patient (Table 2). 14.1% of respondents indicated that workplace culture among surgeons and staff misconceptions regarding SPC is a barrier to SPC consultation for their geriatric trauma patients (Table 2). Meanwhile, 37.5% reported that patient/family misconception of SPC remains a barrier to consultation (Table 2).

**Table 2:**
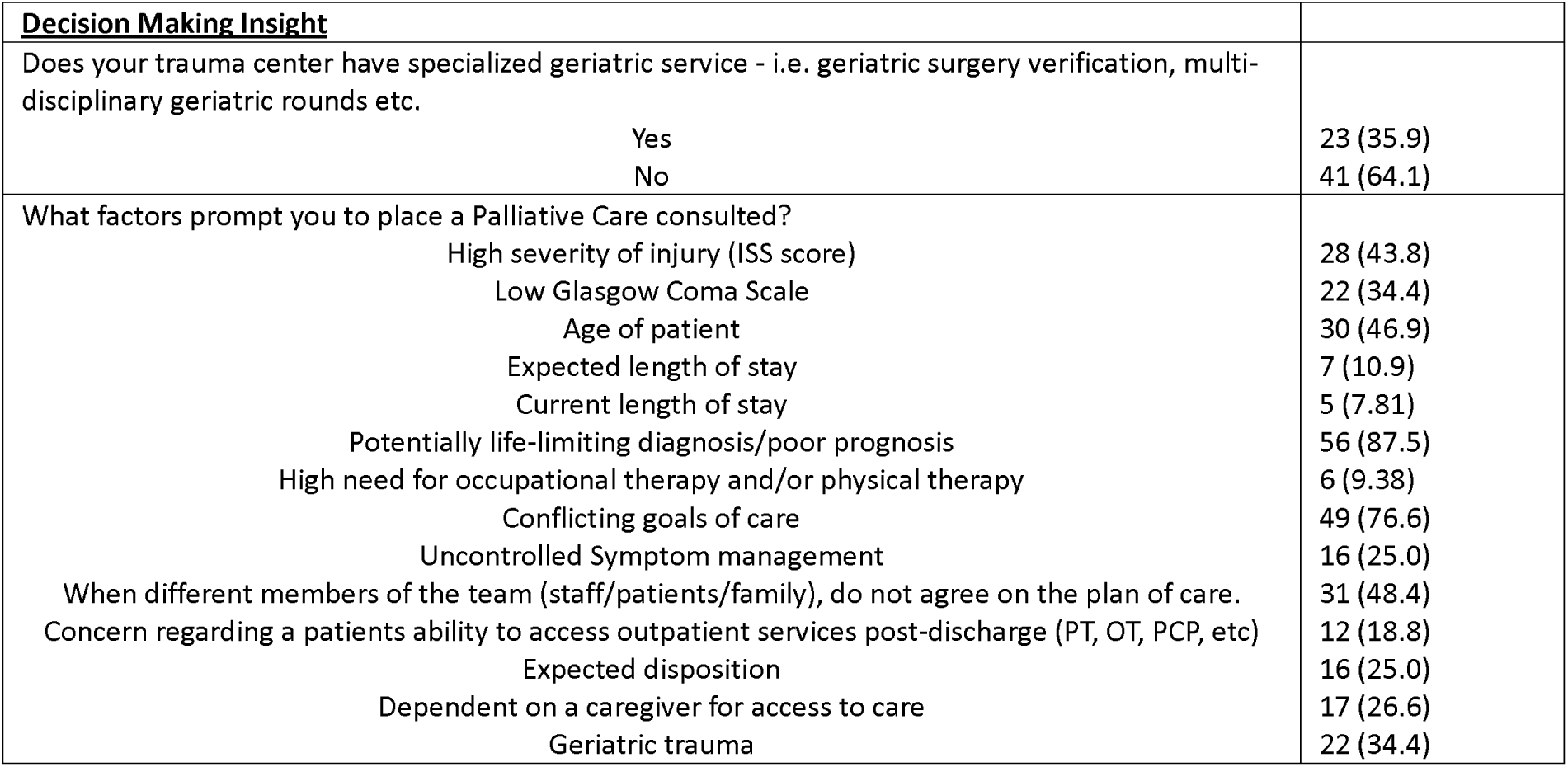
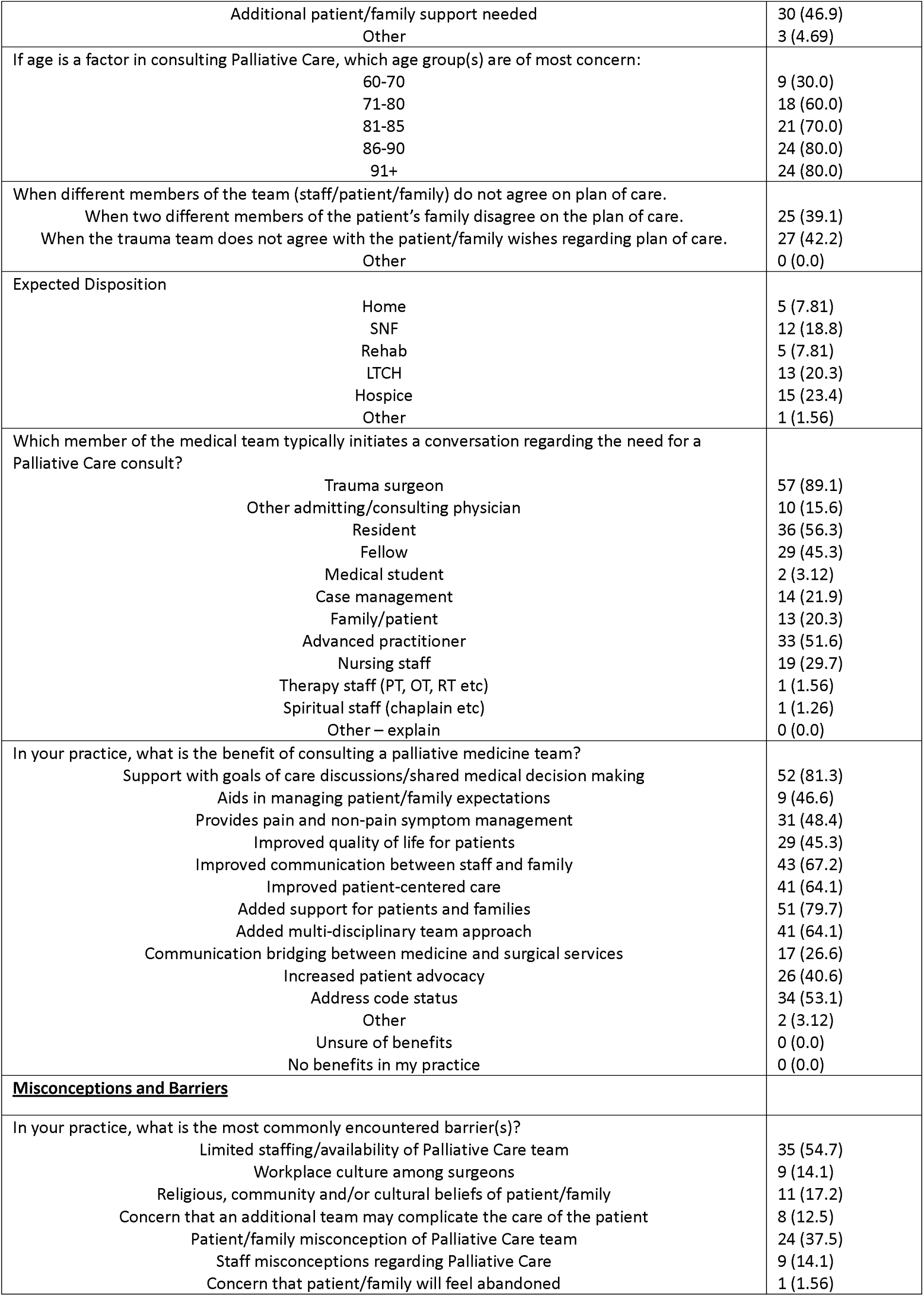
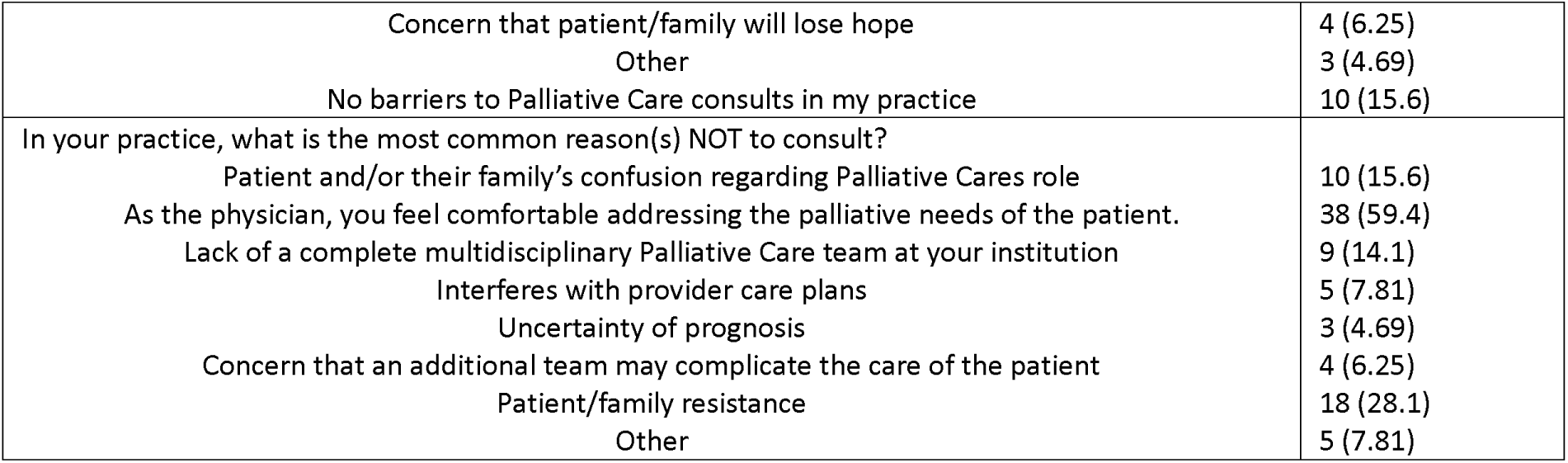
Decision Making Insights and Barriers.

### Physicians’ Stage in Career

Analysis considering years in practice was grouped into early career (residents, fellows and 5 years or less in practice), mid-career (6-15 years of practice), and veteran surgeons (over 15 years in practice) with significant differences regarding SPC consult trends observed across career stages (Table 3). The percentage of respondent who reported “family member disagreement with care plan” as a prompting factor for SPC consult was found to increase with the surgeon’s years in practice (p = 0.017). Additionally, discrepancies between the trauma team and patient/family wishes were the largest among mid-career respondents (p=0.011).

**Table 3.** Decision Making insight and Barriers to Palliative Care stratified by the Physician’s Stage of Career.

| **<u>Decision Making Insight</u>** | **< 5 yrs and**<br>**Resident/Fellow**<br>**N (%)** | **6-15**<br>**N (%)** | **>15 years**<br>**N (%)** | **p-value** |
| --- | --- | --- | --- | --- |
| What factors prompt you to place a Palliative Care consult? |  |  |  |  |
| When different members of the team (staff/patients/family), do not agree on the plan of care. | 8 (38.1) | 12 (75.0) | 10 (41.7) | 0.058 |
| When two different members of the patient's family disagree on the plan of care. | 5 (23.8) | 6 (28.6) | 8 (33.3) | 0.017 |
| When the trauma team does not agree with the patient/family wishes regarding plan of care. | 6 (28.6) | 12 (75.0) | 8 (33.3) | 0.011 |
| Which member of the medical team typically initiates a conversation regarding the need for a Palliative Care consult? |  |  |  |  |
| Resident | 17 (80.9) | 8 (50.0) | 11 (45.8) | 0.043 |
| Fellow | 15 (71.4) | 8 (50.0) | 6 (25.0) | 0.008 |
| Family/patient | 12 (9.52) | 1 (6.25) | 9 (37.5) | 0.023 |
| In your practice, what is the benefit of consulting a palliative medicine team? |  |  |  |  |
| Improved quality of life for patients | 13 (61.9) | 10 (62.5) | 6 (25.0) | 0.018 |
| Increased patient advocacy | 11 (52.4) | 10 (62.5) | 5 (20.8) | 0.017 |
| **<u>Barriers</u>** | **< 5 yrs and**<br>**Resident/Fellow**<br>**N (%)** | **6-15**<br>**N (%)** | **>15 years**<br>**N (%)** | **p-value** |
| In your practice, what is the most commonly encountered barrier(s)? |  |  |  |  |
| Concern that patient/family will lose hope | 0 (0.0) | 4 (25.0) | 0 (0.0) | 0.002 |
| No barriers to palliative care consults in my practice | 6 (28.6) | 3 (18.8) | 1 (4.17) | 0.084 |
| In your practice, what is the most common reason(s) NOT to consult? |  |  |  |  |
| Patient and/or their family's confusion regarding palliative care's role | 1 (4.76) | 5 (31.3) | 4 (16.7) | 0.098 |

Only statistically significant and noticeable trends were reported in this table.

The team members who typically initiate SPC consult considerations were found to be different across the career stages of the attending surgeon. The percentage of residents (p=0.043) and fellows (p=0.008) who initiated SPC discussions decreased as the attending surgeon’s experience level increased. Conversely, for veteran physicians, we found an increase in reported family/patient initiation for SPC (p=0.023). Additionally, veteran surgeon respondents were significantly less likely to identify “increase quality of life” and “increase patient advocacy” as benefits of SPC consultation compared to less experienced surgeons (p=0.018 and p=0.017, respectively).

Barriers in SPC across years in practice had limited significant results. Of clinical interest, reported patient/family confusion regarding SPC role was noticeably different across experience levels, although not statistically significant.

### Setting

Decision-making insight and barriers within the practice settings (urban, academic, and community) are reported in Table 4. There was a statistical difference in the age groups viewed as most needing an SPC consultation. Within urban hospital settings, patients 86 years of age or older are viewed as having a greater need for SPC compared to the other two settings (p=0.026). Additionally, respondents from urban hospital settings reported a higher percentage of addressing code status as a benefit to consulting SPC (p=0.005).

**Table 4.** Decision Making insight and Barriers to Palliative Care Stratified by Hospital Setting.

| **<u>Decision Making Insight</u>** | **Other**<br>**N (%)** | **Urban**<br>**N (%)** | **<u>P-values</u>** |
| --- | --- | --- | --- |
| If age is a factor in consulting palliative care, which age group(s) are of most concern:<br>86-90 | 7 (58.3) | 17 (94.4) | 0.026 |
| 91+ | 7 (58.3) | 17 (94.4) | 0.026 |
| In your practice, what is the benefit of consulting a palliative medicine team?<br>Address code status | 9 (32.1) | 25 (69.4) | 0.005 |
| Other |  | Academic |  |
| Does your trauma center have specialized geriatric service - i.e. geriatric surgery verification, multi-disciplinary geriatric rounds etc. (yes, %) | 2 (10.0) | 21 (47.7) | 0.009 |
| Which member of the medical team typically initiates a conversation regarding the need for a Palliative Care consult?<br>Trauma surgeon | 15 (75.0) | 42 (95.5) | 0.026 |
| In your practice, what is the benefit of consulting a palliative medicine team?<br>Support with goals of care discussions/shared medical decision making | 13 (65.0) | 39 (88.6) | 0.038 |
| Aids in managing patient/family expectations | 11 (55.0) | 38 (86.4) | 0.01 |
| Improved quality of life for patients | 5 (25.0) | 24 (54.6) | 0.033 |
| Improved patient-centered care | 8 (40.0) | 33 (75.0) | 0.011 |
| Added multi-disciplinary team approach | 8 (40.0) | 33 (75.0) | 0.011 |
| Other |  | Community |  |
| What factors prompt you to place a Palliative Care consult?<br>Low Glasgow Coma Scale | 14 (27.5) | 8 (61.5) | 0.046 |
| Dependent on a caregiver for access to care | 10 (19.6) | 7 (53.9) | 0.03 |
| **<u>Barriers</u>** | **Other**<br>**N (%)** | **Urban**<br>**N (%)** | **<u>P-values</u>** |
| In your practice, what is the most common reason(s) NOT to consult?<br>Uncertainty of prognosis | 3 (10.7) | 0 (0.0) | 0.044 |
| Other |  | Academic |  |
| In your practice, what is the most commonly encountered barrier(s)?<br>Limited staffing/availability of palliative care team | 7 (35.0) | 28 (63.4) | 0.033 |
| In your practice, what is the most common reason(s) NOT to consult?<br>As the physician, you feel comfortable addressing the palliative needs of the patient. | 8 (40.0) | 30 (68.2) | 0.033 |
| Other |  | Community |  |
| In your practice, what is the most common reason(s) NOT to consult?<br>Other | 2 (3.92) | 3 (23.1) | 0.022 |

Only statistically significant and noticeable trends were reported in this table.

Trauma surgeons from academic centers reported having specialized geriatric services significantly more often than surgeons from combined urban and community settings (47.7% vs 10.0%; p=0.009). Furthermore, the respondents within the academic centers were the team members who typically initiated the conversation regarding SPC with the patients (p=0.026). Trauma surgeons from academic centers also reported a statistically significant increase in the percentages of specific benefits consulting SPC provides to patients. For example, benefits observed within the academic centers included shared medical decision-making and discussions of patient and provider goals (p=0.038). A higher percentage of surgeons from academic centers believe that the SPC team helps with patient and family expectations compared to the urban and community settings (p=0.01). Other reported perceived benefits include an improved quality of life for the patient, improved patient-centered care, and a multi-disciplinary team approach (p<0.05).

The differences in main barriers were observed between the academic centers and the combined urban/community centers. Limited staffing and the unavailability of an SPC were perceived as the main barriers in academic settings (p=0.033). Furthermore, 68.2% of the respondents within the academic setting felt comfortable addressing the palliative needs of the patients (p=0.033). In addition, 10.7% of the academic and community settings reported that the uncertainty of prognosis was the most common reason not to consult with a SPC.

### Percentage of Geriatrics Seen

Insight and barriers were compared across estimated geriatric percentages in the patient population. The reported geriatric patient percentages were grouped into four tiers: less than or equal to 25%, 26-50%, 51-75%, and over 75%, referred to as low, medium, high, and very high, respectively. (Table 5). The patient’s age as a prompt for SPC consult across percent of geriatric patients was significant (p=0.014). Respondents from hospitals with a greater than 25% geriatric patient population reported a higher percentage of age considerations for their patients.

**Table 5.** Decision Making insight and Barriers to Palliative Care stratified by the Percentage of Geriatrics seen.

| **<u>Decision Making Insight</u>** | **Low Geriatric**<br>**Population**<br>**≥25%**<br>**N (%)** | **Medium Geriatric**<br>**Population**<br>**26-50%**<br>**N (%)** | **High Geriatric**<br>**Population**<br>**51-75%**<br>**N (%)** | **Very high Geriatric**<br>**Population**<br>**>75%**<br>**N (%)** | **p-value** |
| --- | --- | --- | --- | --- | --- |
| What factors prompt you to place a Palliative Care consult? |  |  |  |  |  |
| Age of patient | 0 (0.0) | 20 (64.5) | 7 (36.8) | 3 (50.0) | 0.014 |
| Concern regarding a patients ability to access outpatient services post-discharge (PT, OT, PCP, etc) | 1 (16.7) | 4 (12.9) | 3 (15.8) | 4 (66.7) | 0.044 |
| In your practice, what is the benefit of consulting a palliative medicine team? |  |  |  |  |  |
| Aids in managing patient/family expectations | 2 (33.3) | 25 (80.7) | 17 (89.5) | 5 (83.3) | 0.043 |
| **<u>Barriers</u>** | Less Than or Equal To 25%<br>N (%) | 26-50<br>N (%) | 51-75<br>N (%) | >75<br>N (%) | p-value |
| In your practice, what is the most commonly encountered barrier(s)? |  |  |  |  |  |
| Workplace culture among surgeons | 3 (50.0) | 2 (6.45) | 4 (21.1) | 0 (0.0) | 0.025 |
| Concern that patient/family will feel abandoned | 1 (16.7) | 0 (0.0) | 0 (0.0) | 0 (0.0) | 0.023 |
| In your practice, what is the most common reason(s) NOT to consult? |  |  |  |  |  |
| As the physician, you feel comfortable addressing the palliative needs of the patient. | 3 (50.0) | 15 (48.4) | 16 (84.2) | 4 (66.7) | 0.079 |

Only statistically significant and noticeable trends were reported in this table.

Additionally, 66.7% of the surgeons with very high (over 75%) geriatric populations reported concerns regarding patient’s access to outpatient services (p=0.044). When considering the levels of geriatric patients, patient/family expectations as an SPC consult benefit were significantly identified by respondents (p= 0.043). Workplace culture among surgeons, as a commonly encountered barrier, was significantly different across the levels of geriatric population (p=0.025). Clinically relevant, surgeon comfort in addressing patient palliative needs was not found to be significantly different. Respondents with a high (51-75%) geriatric population reported the highest percentage of physicians comfortable addressing palliative needs (84.2%), followed by surgeons with a very high (over 75%) geriatric practice (66.7%). None of the respondents with a very high geriatric patient population reported workplace culture or patient/family abandonment as barriers.

### Amount Consulted in Palliative Care among Geriatric Trauma Patients

As the amount of Palliative Care consulted increased, the factors that prompted the respondents to place SPC included potentially life-limiting diagnosis or poor prognosis and a geriatric trauma (p<0.05) (Table 6). Furthermore, although not statistically significant, consideration of the patient’s age also increased relative to the physicians’ relative use of SPC consults. Respondents who often/always seek SPC consultations observed the highest percentage of perceived benefits of such intervention. The benefits identified by often/always SPC consulting surgeons included managing family or patient expectations, improved communication between staff and family, and added support for patients and their families. Conversely, surgeons who rarely consult SPC report that a lack of a complete multidisciplinary SPC team is the most common barrier seen at their institutions (p=0.042).

**Table 6.** Decision Making insight and Barriers to Palliative Care stratified by the Likert Amount of Palliative Medicine Consulted.

| **<u>Decision Making Insight</u>** | Rarely<br>N (%) | Sometimes<br>N (%) | Often/Always<br>N (%) | p-value |
| --- | --- | --- | --- | --- |
| What factors prompt you to place a Palliative Care consulted? |  |  |  |  |
| Age of patient | 2 (33.3) | 8 (32.0) | 16 (64.0) | 0.07 |
| Potentially life-limiting diagnosis/poor prognosis | 4 (66.7) | 23 (92.0) | 25 (100.0) | 0.027 |
| Geriatric trauma | 0 (0.0) | 5 (20.0) | 15 (60.0) | 0.02 |
| If age is a factor in consulting palliative care, which age group(s) are of most concern: |  |  |  |  |
| 60-70 | 0 (0.0) | 0 (0.0) | 6 (37.5) | 0.095 |
| 71-80 | 0 (0.0) | 4 (50.0) | 12 (75.0) | 0.090 |
| In your practice, what is the benefit of consulting a palliative medicine team? |  |  |  |  |
| Aids in managing patient/family expectations | 3 (50.0) | 20 (80.0) | 23 (92.0) | 0.054 |
| Improved communication between staff and family | 3 (50.0) | 16 (64.0) | 22 (88.0) | 0.055 |
| Added support for patients and families | 4 (66.7) | 20 (80.0) | 24 (96.0) | 0.087 |
| **<u>Barriers</u>** | Rarely<br>N (%) | Sometimes<br>N (%) | Often/Always<br>N (%) | p-value |
| In your practice, what is the most commonly encountered barrier(s)? |  |  |  |  |
| Patient/family misconception of palliative care team | 0 (0.0) | 13 (52.0) | 8 (32.0) | 0.046 |
| In your practice, what is the most common reason(s) NOT to consult?<br>Lack of a complete multidisciplinary palliative care team at your institution | 3 (50.0) | 4 (16.0) | 2 (8.00) | 0.042 |

## Discussion

Early and effective SPC involvement in geriatric trauma care has a meaningful impact on patient outcomes and quality of life while decreasing healthcare utilization. However, despite best practice guidelines promoting SPC consultation, usage continues to be inconsistent and imprecise, often delivered late or upon uncontrolled symptoms, diminishing the effectiveness. This study contributes new pragmatic knowledge derived from 64 responding trauma surgeons’ perspectives on SPC and the barriers hindering its utilization in geriatric trauma care.

Table 3 displays survey data stratified by physician stage of career. Regarding decision-making insight questions, factors related to interpersonal conflict between family members/patients or patients/families and care teams showed significant differences across career stages (p=0.017, p=0.011, respectively). Similarly, the conflict between members of the care team was near significant. These data suggest that SPC aids communication, especially when considering the physician career stage. Furthermore, residents/fellows and patients/family were reported to initiate SPC considerations significantly more often when with early career attending surgeons compared to more experienced surgeons. Early career surgeon respondents were also more likely to recognize “increased quality of life” and “increased patient advocacy” as SPC benefits than veteran surgeon respondents. However, only mid-career surgeons reported “patient/family to lose hope” as the most common barrier to SPC consult (p=0.002). It is currently unclear why these career stage trends are observed amongst the respondents. However, it is reasonable to consider the growth of SPC as a subspeciality across the healthcare system, and increased non-PC physician and medical student education efforts in the past 15 years may play a role in these observations ^33,34^. American Academy of Hospice and Palliative Medicine, Center to Advance Palliative Care, National Coalition for Hospice and Palliative Care, and other specialty and patient organizations have ongoing national initiatives to increase Hospice and Palliative Care education to learners and practicing physicians alike ^33–36^.

Three interesting trends regarding responding trauma surgeons’ barriers to SPC were identified across the percentage of geriatric patients (table 4). Workplace culture around SPC was most reported as a barrier by surgeons with a reported low geriatric population (p=0.025). Only one respondent with a reported low geriatric patient population indicated “concern of patient/family will feel abandoned” as a barrier to PC. The inverse relationship between this barrier and the frequency of geriatric trauma implies a known benefit of SPC in practices with a high volume of geriatric trauma patients amongst respondents. Consideration of these data may be of value for cross-department professional education efforts within a hospital or the greater healthcare system with demographic similarity to the survey respondents. Also of note, trauma surgeons’ level of comfort addressing the SPC needs of patients was not significantly different across the percentage of geriatric patients seen. Based on this finding, it is reasonable to infer that baseline education in primary SPC needs has become more widely available amongst responding surgeons. Furthermore, this finding supports the general trends of the national expansion of primary PC by the admitting physician and secondary PC, a consultant, since 2000^33,34^.

Significant differences regarding SPC consults prompting factors across the amount of SPC consults were identified (table 5). Differences were identified between the Likert frequency of SPC consultation utilization (rarely, sometimes, often/always) and “geriatric trauma” as the prompting factor (p = 0.02). A similar trend was seen with “life-limiting diagnosis/poor prognosis” (p=0.027). There were no significant differences across SPC consult frequencies regarding the benefits of SPC, which indicates that understanding of SPC benefits in geriatric trauma is unrelated to the frequency of SPC utilization. These data imply that physician knowledge and/or misconceptions alone are not related to utilization amounts. Conversely, a lack of multidisciplinary SPC team access does pose a barrier with significant differences across levels of SPC utilization (p=0.042). Most respondents who identified a lack of access reported SPC consults as “rare” or “sometimes” in geriatric patients. These findings support (1) the general effectiveness of SPC education efforts amongst the responding trauma surgeons and (2) the continued need for primary palliative care education amongst surgeons, especially when access to SPC is limited.

This study had several limitations. Most significantly a poor response rate and the use of display logic decreased the response rate for some items. The only 2.8% response from the AAST memberships for this study limits the understanding of SPC utilization in geriatric trauma patients for all AAST members. Therefore, much of the data and subsequent inferences are limited to interpretation of responses, with great restriction on generalizability. The generalizability of the study is limited by using convenient sampling and repetition of sending survey invitations was our only effort to address potential nonresponse bias. Efforts to make more granular comparisons between practice settings, years in practice, board certification, regularity of SPC consults, and percentage of geriatric patients are limited by selection bias due to the low number of respondents and high representation of level 1 trauma centers. Although response and self-selection bias do influence the data set and subsequent analysis, a majority (78.1%, 50/64, Table 1) of respondents indicated geriatric trauma patients make up between 25-75% of their practice, suggesting that these data represent insights from 64 surgeons well-experienced and expert in geriatric trauma management. Although the survey was created collaboratively across relevant disciplines and with multiple rounds of review, it is not a formally validated tool. Despite these limitations, the study achieved its aims within its own limitations. The results provided insights, not previously reported by small or large samples, from 64 trauma surgeons into their current usage of SPC consults in geriatric trauma and factors contributing to the related decision-making and barriers.

## Conclusion

Few geriatric trauma patients receive the maximum benefit of SPC despite the known benefits of early palliative care and established best-care guidelines. This single-timepoint survey study aimed to elucidate decision-making considerations and barriers responding trauma surgeons face when consulting SPC for geriatric patients. These data indicate a known benefit of SPC amongst responding trauma surgeons and generally support that geriatric trauma is considered an opportunity for SPC collaboration. This study initiated the exploration into practical understanding needed to enhance pragmatic SPC integration in geriatric trauma care.

## Data Availability

All data produced in the present study are available upon reasonable request to the authors

---

Supplementary Figure 1: Survey PDF

## Notes

### Competing Interest Statement

The authors have declared no competing interest.

### Funding Statement

This study did not receive any funding

### Author Declarations

Dignity Health St. Josephs Medical Centers Institutional Review Board (IRB# PHXNR-23-500-253-71-47)

