## supplemental tables for "Palliative Care in Geriatric Trauma: Quantitative Insight From 64 Trauma Surgeon Survey Respondent on Utilizing Specialty Palliative Care in Geriatric Trauma"

Supplemental Table 1. Descriptive statistics of Palliative Care Decision Making Process and Perceived Barriers to Palliative Care.

| **Decision Making Insight** |  |
| --- | --- |
| Does your trauma center have specialized geriatric service - i.e. geriatric surgery verification, multi-disciplinary geriatric rounds etc. (yes, %) | 23 (35.9) |
| What factors prompt you to place a Palliative Care consulted?  High severity of injury (ISS score)  Low Glasgow Coma Scale  Age of patient  Expected length of stay  Current length of stay  Potentially life-limiting diagnosis/poor prognosis  High need for occupational therapy and/or physical therapy  Conflicting goals of care  Uncontrolled Symptom management  When different members of the team (staff/patients/family), do not agree on the plan of care.  Concern regarding a patients ability to access outpatient services post-discharge (PT, OT, PCP, etc)  Expected disposition  Dependent on a caregiver for access to care  Geriatric trauma  Additional patient/family support needed  Other | 28 (43.8)  22 (34.4)  30 (46.9)  7 (10.9)  5 (7.81)  56 (87.5)  6 (9.38)  49 (76.6)  16 (25.0)  31 (48.4)  12 (18.8)  16 (25.0)  17 (26.6)  22 (34.4)  30 (46.9)  3 (4.69) |
| If age is a factor in consulting palliative care, which age group(s) are of most concern:  60-70  71-80  81-85  86-90  91+ | 9 (30.0)  18 (60.0)  21 (70.0)  24 (80.0)  24 (80.0) |
| When different members of the team (staff/patient/family) do not agree on plan of care.  When two different members of the patient’s family disagree on the plan of care.  When the trauma team does not agree with the patient/family wishes regarding plan of care.  Other | 25 (39.1)  27 (42.2)  0 (0.0) |
| Expected Disposition  Home  SNF  Rehab  LTCH  Hospice  Other | 5 (7.81)  12 (18.8)  5 (7.81)  13 (20.3)  15 (23.4)  1 (1.56) |
| Which member of the medical team typically initiates a conversation regarding the need for a Palliative Care consult?  Trauma surgeon  Other admitting/consulting physician  Resident  Fellow  Medical student  Case management  Family/patient  Advanced practitioner  Nursing staff  Therapy staff (PT, OT, RT etc)  Spiritual staff (chaplain etc)  Other – explain | 57 (89.1)  10 (15.6)  36 (56.3)  29 (45.3)  2 (3.12)  14 (21.9)  13 (20.3)  33 (51.6)  19 (29.7)  1 (1.56)  1 (1.26)  0 (0.0) |
| In your practice, what is the benefit of consulting a palliative medicine team?  Support with goals of care discussions/shared medical decision making  Aids in managing patient/family expectations  Provides pain and non-pain symptom management  Improved quality of life for patients  Improved communication between staff and family  Improved patient-centered care  Added support for patients and families  Added multi-disciplinary team approach  Communication bridging between medicine and surgical services  Increased patient advocacy  Address code status  Other  Unsure of benefits  No benefits in my practice | 52 (81.3)  9 (46.6)  31 (48.4)  29 (45.3)  43 (67.2)  41 (64.1)  51 (79.7)  41 (64.1)  17 (26.6)  26 (40.6)  34 (53.1)  2 (3.12)  0 (0.0)  0 (0.0) |

Supplemental Table 2. Descriptive statistics of Perceived Barriers to Palliative Care.

| **Misconceptions and Barriers** |  |
| --- | --- |
| In your practice, what is the most commonly encountered barrier(s)?  Limited staffing/availability of palliative care team  Workplace culture among surgeons  Religious, community and/or cultural beliefs of patient/family  Concern that an additional team may complicate the care of the patient  Patient/family misconception of palliative care team  Staff misconceptions regarding palliative care  Concern that patient/family will feel abandoned  Concern that patient/family will lose hope  Other  No barriers to palliative care consults in my practice | 35 (54.7)  9 (14.1)  11 (17.2)  8 (12.5)  24 (37.5)  9 (14.1)  1 (1.56)  4 (6.25)  3 (4.69)  10 (15.6) |
| In your practice, what is the most common reason(s) NOT to consult?  Patient and/or their familys confusion regarding palliative cares role  As the physician, you feel comfortable addressing the palliative needs of the patient.  Lack of a complete multidisciplinary palliative care team at your institution  Interferes with provider care plans  Uncertainty of prognosis  Concern that an additional team may complicate the care of the patient  Patient/family resistance  Other | 10 (15.6)  38 (59.4)  9 (14.1)  5 (7.81)  3 (4.69)  4 (6.25)  18 (28.1)  5 (7.81) |

Supplemental Table 3. Decision Making insight to Palliative Care stratified by the Board Certification.

| **Decision Making Insight** |  |  |  |
| --- | --- | --- | --- |
|  | Other | Trauma and acute care surgery | **P-values** |
|  | N (%) | N (%) |  |
| What factors prompt you to place a Palliative Care consulted?  Conflicting goals of care | 7 (16.7) | 9 (40.9) | 0.066 |
| Which member of the medical team typically initiates a conversation regarding the need for a Palliative Care consult?  Resident  Nursing staff | 20 (47.6)  16 (38.1) | 16 (72.7)  3 (13.6) | 0.067  0.049 |
| In your practice, what is the benefit of consulting a palliative medicine team?  Communication bridging between medicine and surgical services | 17 (40.5) | 0 ( 0.0) | <0.001 |
|  | Other | General surgery |  |
| What factors prompt you to place a Palliative Care consulted?  Age of patient  Additional patient/family support needed | 2 (20.0)  1 (10.0) | 28 (51.9)  29 (53.7) | 0.088  0..015 |
| Which member of the medical team typically initiates a conversation regarding the need for a Palliative Care consult?  Trauma surgeon | 6 (60.0) | 51 (94.4) | 0.009 |
| In your practice, what is the benefit of consulting a palliative medicine team?  Support with goals of care discussions/shared medical decision making  Aids in managing patient/family expectations  Improved communication between staff and family  Improved patient-centered care  Added support for patients and families  Added multi-disciplinary team approach  Address code status | 6 (60.0)  3 (30.0)  4 (40.0)  3 (30.0)  4 (40.0)  3 (30.0)  2 (20.0) | 46 (85.2)  46 (85.2)  39 (72.2)  38 (70.4)  47 (87.0)  38 (70.4)  32 (59.3) | 0.082  <0.001  0.068  0..028  0.003  0.028  0.036 |
|  | Other | Critical care |  |
| Does your trauma center have specialized geriatric service - i.e. geriatric surgery verification, multi-disciplinary geriatric rounds etc. (yes, %) | 4 (25.0) | 19 (39.6) | 0.017 |
| What factors prompt you to place a Palliative Care consulted?  High severity of injury (ISS score)  Low Glasgow Coma Scale  Conflicting goals of care  Expected disposition  Dependent on a caregiver for access to care  Additional patient/family support needed | 3 (18.8)  1 (6.25)  9 (56.3)  1 (6.25)  1 (6.25)  4 (25.0) | 25 (52.1)  21 (43.8)  40 (83.3)  15 (31.3)  16 (33.3)  26 (54.2) | 0.023  0.006  0.041  0.052  0.048  0.5 |
| Expected Disposition  Hospice | 1 (6.25) | 14 (29.2) | 0.089 |
| In your practice, what is the benefit of consulting a palliative medicine team?  Aids in managing patient/family expectations  Added support for patients and families  Address code status | 9 (56.3)  9 (56.3)  5 (31.3) | 40 (83.3)  42 (87.5)  29 (60.4) | 0.041  0.013  0.081 |

Supplemental Table 4. Misconceptions and Barriers to Palliative Care stratified by the Board Certification

| **Barriers** |  |  |  |
| --- | --- | --- | --- |
|  | Other | Trauma and acute care surgery | **P-values** |
|  | N (%) | N (%) |  |
| In your practice, what is the most common reason(s) NOT to consult?  Patient and/or their familys confusion regarding palliative cares role | 9 (21.4) | 1 ( 4.55) | 0.077 |
|  | Other | General surgery |  |
| In your practice, what is the most commonly encountered barrier(s)?  Limited staffing/availability of palliative care team | 2 (20.0) | 33 (61.1) | 0.016 |
| In your practice, what is the most common reason(s) NOT to consult?  As the physician, you feel comfortable addressing the palliative needs of the patient. | 2 (20.0) | 36 (66.7) | 0.006 |
|  | Other | Critical care |  |
| In your practice, what is the most commonly encountered barrier(s)?  Patient/family misconception of palliative care team | 2 (12.5) | 22 (45.8) | 0.017 |
