## supplemental figure 1 for "Palliative Care in Geriatric Trauma: Quantitative Insight From 64 Trauma Surgeon Survey Respondent on Utilizing Specialty Palliative Care in Geriatric Trauma"

Supplemental Figure 1. Full Survey Instrument.



### Utilization of Palliative Care Consult in Geriatric Trauma Care

Please complete the survey below.



#### Demographics

| Question | Response |
| --- | --- |
| Please mark all current or past board certification | ☐ Trauma and Acute Care Surgery<br>☐ General Surgery<br>☐ Critical Care<br>☐ Neurocritical Care<br>☐ Hospice and Palliative Medicine<br>☐ Other |
| Other | ______________________________ |
| How many years have you been a practicing attending physician? | ◯ Currently in residency/fellowship<br>◯ Under 5 years<br>◯ 6-10 years<br>◯ 11-15 years<br>◯ 16-20 years<br>◯ 21+ years |
| Gender | ☐ Female<br>☐ Male<br>☐ Non-binary/third gender<br>☐ Transgender<br>☐ Cisgender<br>☐ Agender<br>☐ Prefer to self-describe<br>☐ Prefer not to say<br>☐ Genderqueer<br>☐ A gender not listed |
| Prefer to self-describe | ______________________________ |
| How would you describe your primary practice setting? Mark all that apply. | ☐ Rural<br>☐ Urban<br>☐ Academic<br>☐ Community |
| What level trauma center do you primarily work at? | ☐ 1<br>☐ 2<br>☐ 3<br>☐ 4<br>☐ Other<br>☐ No sure |
| Explain | ______________________________ |
| Estimate the percentage of geriatric (60+ years old) trauma patients in your current practice. | ◯ Less than 25%<br>◯ 25-50%<br>◯ 51-75%<br>◯ Over 75% |

| | |
|---|---|
| The trauma service at the hospital that I primarily work is a/an | ◯ Admitting service<br>◯ Consulting service |
| Which specialty acts as the admitting service? Mark all that apply | ☐ Internal Medicine/Hospitalist<br>☐ General Surgery<br>☐ Critical Care (internal medicine trained)<br>☐ Critical care (surgery trained)<br>☐ Other |
| Explain | __________________________________ |
| Does your trauma center have a physician run subspeciality Palliative Care consult team? | ◯ Yes<br>◯ No<br>◯ Unsure<br>◯ Other |
| Explain | __________________________________________ |
| How often do you consult Palliative Care in geriatric (60+ years old) trauma patients? | ◯ Never<br>◯ Rarely<br>◯ Sometimes<br>◯ Often<br>◯ Always<br>◯ Not applicable |

### Decision Making Insight

Please complete the survey below.

page 2 of 3

---

Does your trauma center have specialized geriatric service - i.e. geriatric surgery verification, multi-disciplinary geriatric rounds etc.

- ◯ Yes
- ◯ No
- ◯ Unsure

---

What factors indicate to YOU a need for a palliative care consultation? Check all

- ☐ High severity of injury (ISS score)
- ☐ Low Glasgow Coma Scale
- ☐ Age of patient
- ☐ Expected length of stay
- ☐ Current length of stay
- ☐ Potentially life-limiting diagnosis/poor prognosis
- ☐ High need for occupational therapy and/or physical therapy
- ☐ Conflicting goals of care
- ☐ Uncontrolled Symptom management
- ☐ When different members of the team (staff/patients/family), do not agree on the plan of care
- ☐ Concern regarding a patient's ability to access outpatient services post-discharge (PT, OT, PCP, etc)
- ☐ Expected disposition
- ☐ Dependent on a caregiver for access to care
- ☐ Geriatric trauma
- ☐ additional patient/family support needed
- ☐ Other

---

Explain

______________________________________

---

If age is a factor in consulting palliative care, which age group(s) are of most concern (mark all that apply)

- ☐ 60-70 years old
- ☐ 71-80 years old
- ☐ 81-85 years old
- ☐ 86-90 years old
- ☐ 91+ years old

---

Expected Length of Stay: Explain

______________________________________

---

Current Length of Stay: Explain

______________________________________

---

Symptom Management: Explain

______________________________________

---

When Members of the Team or Family Disagree

- ☐ When two different members of the patient's family disagree on the plan of care.
- ☐ When the trauma team does not agree with the patient/family's wishes regarding the plan of care.
- ☐ Other

Explain

______________________________

Expected Disposition (Mark all that apply)

- ☐ Home
- ☐ SNF
- ☐ Rehab
- ☐ LTCH
- ☐ Hospice
- ☐ Other

Explain

______________________________

Which member of the medical team typically initiates a conversation regarding the need for a Palliative Care consult? Mark all that apply

- ☐ Trauma surgeon
- ☐ Other admitting/consulting physician
- ☐ Resident
- ☐ Fellow
- ☐ Medical student
- ☐ Case management
- ☐ Family/patient
- ☐ Advanced practitioner
- ☐ Nursing staff
- ☐ Therapy staff (PT, OT, RT etc)
- ☐ Spiritual staff (chaplain etc)
- ☐ Other

Explain

______________________________

### Palliative Care Use

Please complete the survey below.

Page 3 of 3

| | |
|---|---|
| In your practice, what is the benefit of consulting a palliative medicine team? Mark all that apply | ☐ Support with goals of care discussions/shared medical decision making<br>☐ Aids in managing patient/family expectations<br>☐ Provides pain and non-pain symptom management<br>☐ Improved quality of life for patients<br>☐ Improved communication between staff and family<br>☐ Improved patient-centered care<br>☐ Added support for patients and families<br>☐ Added multi-disciplinary team approach<br>☐ Communication bridging between medicine and surgical services<br>☐ Increased patient advocacy<br>☐ Address code status<br>☐ Other<br>☐ Unsure of benefits<br>☐ No benefits in my practice |
| Benefits of palliative care: other | ________________________ |
| In your practice, what is the most commonly encountered barrier(s)? Mark all that apply | ☐ Limited staffing/availability of palliative care team<br>☐ Workplace culture among surgeons<br>☐ Religious, community and/or cultural beliefs of patient/family<br>☐ Concern that an additional team may complicate the care of the patient<br>☐ Patient/family misconception of palliative care team<br>☐ Staff misconceptions regarding palliative care<br>☐ Concern that patient/family will feel abandoned<br>☐ Concern that patient/family will lose hope<br>☐ Other<br>☐ No barriers to palliative care consults in my practice |
| Barriers to palliative care consult: other | ________________________ |
| In your practice, what is the most common reason(s) NOT to consult? Mark all that apply | ☐ Patient and/or their family's confusion regarding palliative care's role<br>☐ As the physician, you feel comfortable addressing the palliative needs of the patient.<br>☐ Lack of a complete multidisciplinary palliative care team at your institution<br>☐ Interferes with provider care plans<br>☐ Uncertainty of prognosis<br>☐ Concern that an additional team may complicate the care of the patient<br>☐ Patient/family resistance<br>☐ Other |

The most common reason to NOT consults palliative:
Other
